# A randomised trial of three face-washing methods for the removal of *Chlamydia trachomatis* from the faces of children with severe active trachoma

**DOI:** 10.64898/2025.12.04.25341197

**Authors:** Katie Greenland, Robert Butcher, Edao Sinba Etu, Wake Abebe, Mesfin Bekele, Gebeyehu Dumessa, Yohannes Sitotaw Addisie, Amen Tessema Ariya, Anthony W. Solomon, Alexandra Czerniewska, David Macleod, Anna Last, Oumer Shafi Abdurahman, Matthew Burton, Aida Abashawl, Wondu Alemayehu

## Abstract

**Objectives:** A three-arm, open, parallel-group randomised trial compared three face-washing methods for cleaning *Chlamydia trachomatis (Ct)* from the faces of children with severe active trachoma. The impact of face-washing on *Ct* on the hands of children and their caregivers, and *Ct* duration on faces and hands were investigated as secondary objectives.

**Methods:** Children aged 1–7 years in Oromia, Ethiopia, were screened for active trachoma. We aimed to recruit 470 children with severe conjunctival inflammation; 141 were expected to have concurrent conjunctival infection and facial *Ct*, detected by qPCR. Those with severe inflammation were randomly assigned to water alone, water with soap, or damp, microfibre towel protocols. Swabs (children’s faces/hands, caregivers’ hands), were collected pre-wash, post-wash, and at one, two, four, six and eight hours. Conjunctival swabs were tested for ocular *Ct* infection; only infected children with facial *Ct* were included in primary analysis, comparing the proportion of faces without *Ct* after each washing method.

**Results:** Of 470 children screened with severe inflammation, 25 (5%) had conjunctival infection and facial *Ct*. All three protocols (n=12 water only, n=8 water and soap, n=5 damp towel) reduced discharge, but none removed *Ct* from faces immediately post-wash. No major facial *Ct* load reduction was observed. Face-washing removed *Ct* from some children’s and caregivers’ hands, but loads were not significantly reduced where *Ct* persisted. Though *Ct* was transiently absent from one face at 1 hour and four at 2 hours post-wash, all baseline *Ct*-positive children retained it at eight hours.

**Conclusions:** No evidence of differential or overall effectiveness of the three washing methods at removing facial *Ct* from children with trachoma was found. This finding is limited by a smaller-than-anticipated sample size, potentially hindering detection of subtle differences or overall effects.

**Trial registration:** The trial is registered on ISRCTN (ISRCTN12814010).

## Background

Trachoma, an important infectious cause of blindness, results from conjunctival infection with *Chlamydia trachomatis* (*Ct*) and is targeted for global elimination (1). The World Health Organization recommends the SAFE strategy (surgery, antibiotics, facial cleanliness, environmental improvement). The antibiotic mass drug administration (MDA) (A component) aims to clear conjunctival *Ct* infection (2). A key elimination criterion is sustained prevalence below 5% for trachomatous inflammation—follicular (TF) in 1–9-year-olds for two years without MDA (3). Repeated *Ct* infection cycles are thought to cause blinding complications (4,5), with sustained reductions in *Ct* transmission intensity believed to prevent disease progression.

During *Ct* infection, viable elementary bodies release into ocular and nasal secretions (6–9). Transmission is thought to primarily occur directly on skin and via flies and fomites (10), making frequent face-washing crucial for disruption (11). A primary goal of facial cleanliness (F) interventions is reducing facial *Ct* in children, thereby limiting reinfection and transmission. A potential secondary benefit of face-washing is indirectly reducing *Ct* on hands, further curbing spread.

Despite the strong theoretical basis for facial cleanliness in trachoma control, limited evidence guides effective intervention development (12,13). Specifically, data on optimal face-washing for maximum impact are scarce (14). While water availability, sociocultural factors and lack of agreed-upon facial cleanliness indicators (15,16) complicate development of global guidance on face-washing (17,18), empirical data on core technical components (e.g., soap use, wash cloth, duration and intensity of wash), are essential to focus intervention design (19).

Evaluating facial cleanliness interventions faces methodological challenges. First, active trachoma causes ocular discharge, which can confound the relationship between face-washing, cleanliness and trachoma reduction. Second, self-reported, direct observed behaviours and proxies for behaviour are prone to bias (15,16). Third, facial cleanliness is a transient marker varying throughout the day (20). Consequently, evidence for face-washing’s protective effect against trachoma is mixed (21–26).

Here, our primary objective was to compare the effectiveness of three washing modalities at removing *Ct* from faces of children with trachoma. Secondary objectives investigated effect persistence and the indirect impact on *Ct* carriage on children’s or caregivers’ hands.

## Methods

### Recruitment strategy

Children with conjunctival infection and facial *Ct* were targeted for enrolment in this trial. Central laboratory PCR results for infection had a longer turnaround time than the median duration of *Ct* infection (27). Due to lack of a field-ready point-of-care test, a clinical screening approach was employed to identify children likely to have *Ct.* Participants with severe trachomatous inflammation were enrolled, undergoing ocular and facial *Ct* testing, face-washing and follow-up. Swabs were tested after trial completion; data from those without concurrent conjunctival infection and facial *Ct* were retrospectively excluded from primary analysis. All participants were eligible for secondary analyses.

### Screening for clinical signs of trachoma

The core study team reviewed programmatic trachoma survey data from *woredas* (districts) in Oromia Region to identify potential study sites with a focus on security, lack of recent MDA (past three years), and persistently high or increased TF prevalence in 1–9-year-olds. Five *woredas* were identified in Arsi zone. Community (*kebele*)-level data from the most recent trachoma survey (28) identified communities with the highest TF and/or trachomatous inflammation—intense (TI) prevalence for trial screening.

Teams established a central screening centre in each *kebele*. Community health workers notified residents and encouraged attendance. Home visits were conducted for those unable to travel.

Screening staff were Tropical Data-certified trachoma graders (29,30) who received further training in the 1981 modified WHO (“FPC”) trachoma grading system, which focuses on intensity of visible pathology (31). A hybrid definition of severe trachoma (described below) combined facets of both grading schemes. Graders achieved a kappa score of 0.7 in the grading of TF against an experienced grader, with ongoing periodic review from senior staff.

Children aged 1–7 years (selected for their higher infection risk (27)) were screened. All children with TF and/or TI received tetracycline eye ointment, administered either immediately after screening for ineligible individuals or upon trial exit for enrolled individuals.

### Enrolment in the clinical trial

Based on the positive predictive value of severe follicular or papillary inflammation for ocular *Ct* infection observed elsewhere (9,32,33), children with severe active trachoma were eligible. This was defined as TI (equivalent to P3 in FPC) or ≥10 follicles in the central tarsal conjunctiva (a severe subset of TF, broadly F3 in FPC), in one or both eyes. Participants with ocular and/or facial injury were excluded. After informed consent, enrolled participants returned to the screening centre within 72 hours of screening for all trial activities. Participants were asked to avoid face-washing on the trial morning.

Participants were randomly allocated (1:1:1) to one of three trial arms just before the washing protocol. Allocation was automatically generated by ODK software and displayed on the device. Infection status was unknown at randomisation. The trial was unmasked and recruited all three arms in parallel.

Sample size calculations, informed by our pilot study (19) (33% clearance with water-only wash, assumed least effective), aimed for 80% power to detect a 30 percentage point difference with Bonferroni’s correction (overall 95% confidence, individual comparison confidence ≈ 98.4%). This required 47 children with concurrent conjunctival *Ct* infection and facial *Ct* per arm (141 total). Anticipating a 30% prevalence of conjunctival *Ct* infection among severe trachoma cases (9,32,33), 470 participants were required to achieve the sample size, with recruitment continuing until the target was met.

### Face-washing protocols

Caregivers washed children’s faces using three methods: water alone, water with soap, or a damp microfibre cloth, SuperTowel (Elhra, UK). The SuperTowel is treated with a permanent antimicrobial bonding (quaternary ammonium silane) that kills pathogens upon contact, but does not contain a soluble cleaning agent (34). For water-only and water-and-soap protocols, caregivers washed faces for approximately 30 seconds using study-provided water, with faces and hands air-dried. SuperTowel users (towel-wipe arm) dampened it and wiped faces for approximately 15 seconds; as the SuperTowel was considered an investigational product in the context of this trial, it was collected post-use and disposed of. Locally available, unscented soap bars were provided to the water-with-soap arm at study start, and to other arms after follow-up visits.

Participants returned to the screening centre at one, two, four, six and eight hours post-wash to assess changes in facial *Ct* presence or load over the day. At each visit, caregivers reported any subsequent face-washing.

All protocols were expected to be low risk and well tolerated. Theoretical risks (e.g., soap in the eye, SuperTowel reaction) were acknowledged. Participants were monitored for adverse events during washes and all follow-up visits. Study team contact details were left for post-study reporting.

### Specimen collection

Ocular swabs were collected from each eye by passing a polyester-coated cotton swab across the tarsal conjunctiva thrice, rotating 120° between passes. Face and hand swabs were collected using sterile water-dampened swabs passed over the face and under both eyes and nose, and across finger pads, palms and backs of both hands, respectively.

Ocular swabs were collected pre-wash from every participant. Face and hand swabs (from children and caregivers) were collected immediately before and after the face wash, and at one, two, four, six and eight hours post-wash.

All 940 ocular swabs (both eyes) and 470 pre-wash face swabs were processed to define the primary outcome analysis population. Other face and hand swabs were processed only from children with ocular infection and/or facial *Ct* pre-wash. A random sample of 10 individuals without ocular infection or pre-wash facial *Ct* had all their swabs processed as background controls.

### Laboratory testing

Swabs were immediately placed into coolers after collection and transported for *Ct* testing to Adama Regional Referral and Public Health Laboratory (8,9,32,35), either within one day (districts near to Adama) or weekly after interim cold storage (districts further from Adama). Upon arrival at the laboratory, samples were frozen until processing. DNA was purified from all swabs using the BioChain Blood and Serum DNA Isolation Kit and eluted in 80 uL water. This eluate was then tested for *Homo sapiens RPP30* (endogenous control target), *Ct* outer membrane complex B (*omcB)* and *Ct* plasmid open reading frame2 (*pORF2)* using a published open-platform qPCR assay (35) run on a Thermo Fisher Scientific QuantStudio 7 Flex platform in fast mode. Oligonucleotide concentrations were optimised for this platform. 20-µL assays were made up of TaqMan Multiplex Master Mix, all three primer pairs at 300 nM each, the *H. sapiens RPP30*, *Ct omcB* and *Ct pORF2* probes at 150 nM, 500 nM and 300 nM, respectively, and 4 µL sample eluate. Each sample was tested in one well. On each plate, no-template reactions and a six-step ten-fold dilution series of known-concentration positive control material were tested in two wells each. Thermocycling conditions were 95°C for 20 seconds, followed by 40 cycles of dissociation at 95°C for 1 second and annealing/extension at 60°C for 20 seconds. DNA copy number was extrapolated from the standard curve using a linear equation.

### Assessment of facial cleanliness

Facial cleanliness was assessed by the field team immediately pre-wash and at each post-wash follow up, observing for visible ocular or nasal discharge under natural daylight (36). Two graders assessed and agreed upon facial cleanliness markers in the field. Graders were standardised against grades assigned by an experienced trainer, with retraining as needed until competence was achieved (chance-corrected agreement (Cohen’s kappa) of ≥ 0.90 on all indicators.

Smartphone photographs (Google Pixel 5, 12MP rear camera), of participants’ faces (forehead to mouth) were taken pre- and immediately post-wash to assess cleaning effectiveness. Paired photographs were displayed side-by-side, with left-to-right order randomised (https://chrissyhroberts.github.io/Code_Book/Join_photos_randomly.html). Two masked expert graders assessed cleanliness equivalence, focusing on discharge and dirt, and disregarding flies. Disagreements were resolved by discussion. Face photographs were also collected for repeat grading by the field team and for external review. Facial cleanliness was measured using the quantitative personal hygiene assessment tool (15) at all timepoints. Caregivers provided household water and sanitation access information.

This report focuses on pre- and immediately post-wash visual inspection of facial cleanliness in the field and photographs graded by external graders, with the latter used to demonstrate challenges in assessing cleanliness solely via field observations of discharge. Analysis of household water access and other facial cleanliness data will be presented in future manuscripts.

### Data analysis

The primary analysis focused on participants with PCR-confirmed conjunctival *Ct* infection (at least one eye positive for one of two *Ct* targets and human endogenous control within 40 cycles). PCR results were quantified by extrapolating the quantitation cycle to a standard *Ct* DNA curve. Individual infection load was defined by the eye with the highest *Ct* load.

The primary outcome was the comparison of the effectiveness of face washing with water and soap versus face washing with water only at removing *Ct* from faces. This effectiveness was determined as the difference between the proportion of faces without *Ct* post-wash among those with baseline conjunctival infection and facial *Ct* in the two groups. The effectiveness of face washing with water alone and face washing with water and soap were also both compared to face washing with SuperTowel (secondary outcomes). These between-group differences in proportions (with 95% CI) were reported for each washing method and compared using pairwise Fisher’s exact tests. The same method applied to the secondary outcome of field-graded oculo-nasal discharge. To estimate the wash’s effectiveness at reducing facial *Ct* load (secondary outcome), individual *Ct* load changes (pre-wash load minus post-wash load) were calculated and compared between groups using a Mann-Whitney U test.

Face wash effectiveness in removing *Ct* from children’s or caregivers’ hands (*ad hoc* secondary outcomes) was determined by the change in proportion of contaminated hands post-wash. Similar to facial *Ct* outcomes, Fisher’s exact tests and Mann Whitney U tests assessed significance for binomial and continuous outcomes, respectively.

No prolonged protective effect was expected from the wash protocols. Thus, protocol-specific analyses of marker changes over the day were not conducted. Instead, marker changes were compared between those with and without conjunctival *Ct* infection.

## Results

### Screening and enrolment

Screening, recruitment and trial participation occurred between April and July 2023, just before the rainy season. In 43 *kebeles* across five *woredas* in Arsi Zone, Oromia region, 14,716 children aged 1–7 years were screened. Of these, 1,694 (12%) had TF and 40 (<1%) had TI. For monitoring purposes, each grade was counted without distinguishing laterality, differing severity, or multiple signs in one eye. Among those with TF, 450 (27%) had ≥10 follicles and were defined as severe cases.

From the 490 eligible children, 470 children with severe active trachoma were enrolled. Of these, 422 (90%) had severe active trachoma in both eyes, while the remaining 48 (10%) had mild TF (5–9 central follicles) in one eye and severe active trachoma in the other. Ten out of 470 (2%) individuals had both ≥10 follicles and TI in at least one eye. Randomisation allocated 170 (36%) to the water-only arm, 150 (32%) to the water-and-soap arm, and 150 (32%) to the towel-wipe arm (Figure 1).

**Figure 1.**
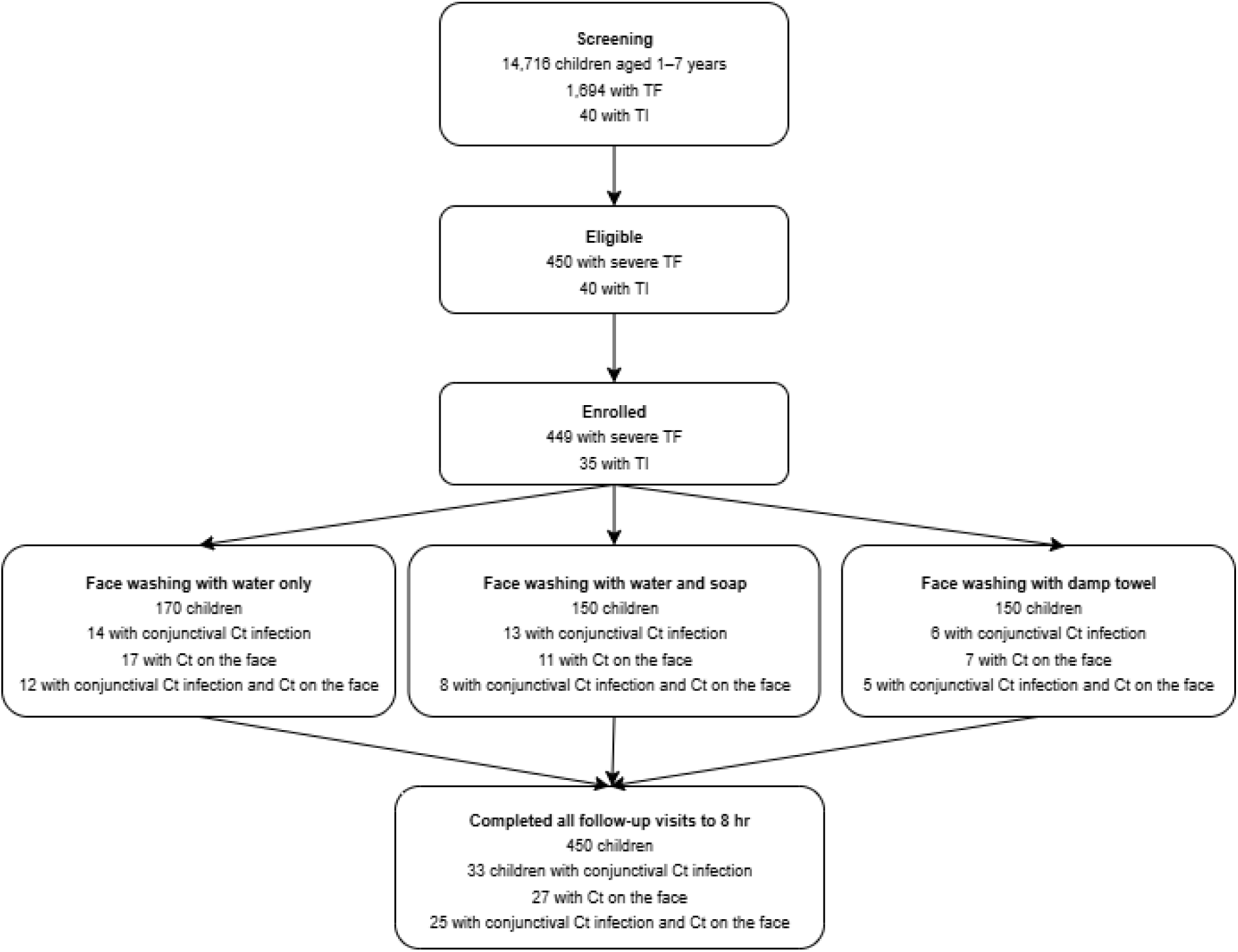
Trial profile.

Of the 470 enrolled, 33 (7%) had conjunctival *Ct* infection in at least one eye, with 25/33 (76%) infected children having bilateral infection. Infected participants were from 16 households across four *kebeles*. The median conjunctival infection load was 14,083 *omcB* copies/µL (range: 1,563–64,051 *omcB* copies/µL).

25/33 (76%) children with conjunctival infection also had *Ct* on their face. Additionally, 10 children had facial *Ct* without conjunctival infection. Per prospective inclusion criteria (primary analysis restricted to children with both conjunctival and facial *Ct*), 445 children were excluded. Participant characteristics for the primary analysis are shown in Table 1. No adverse events were reported.

**Table 1.**
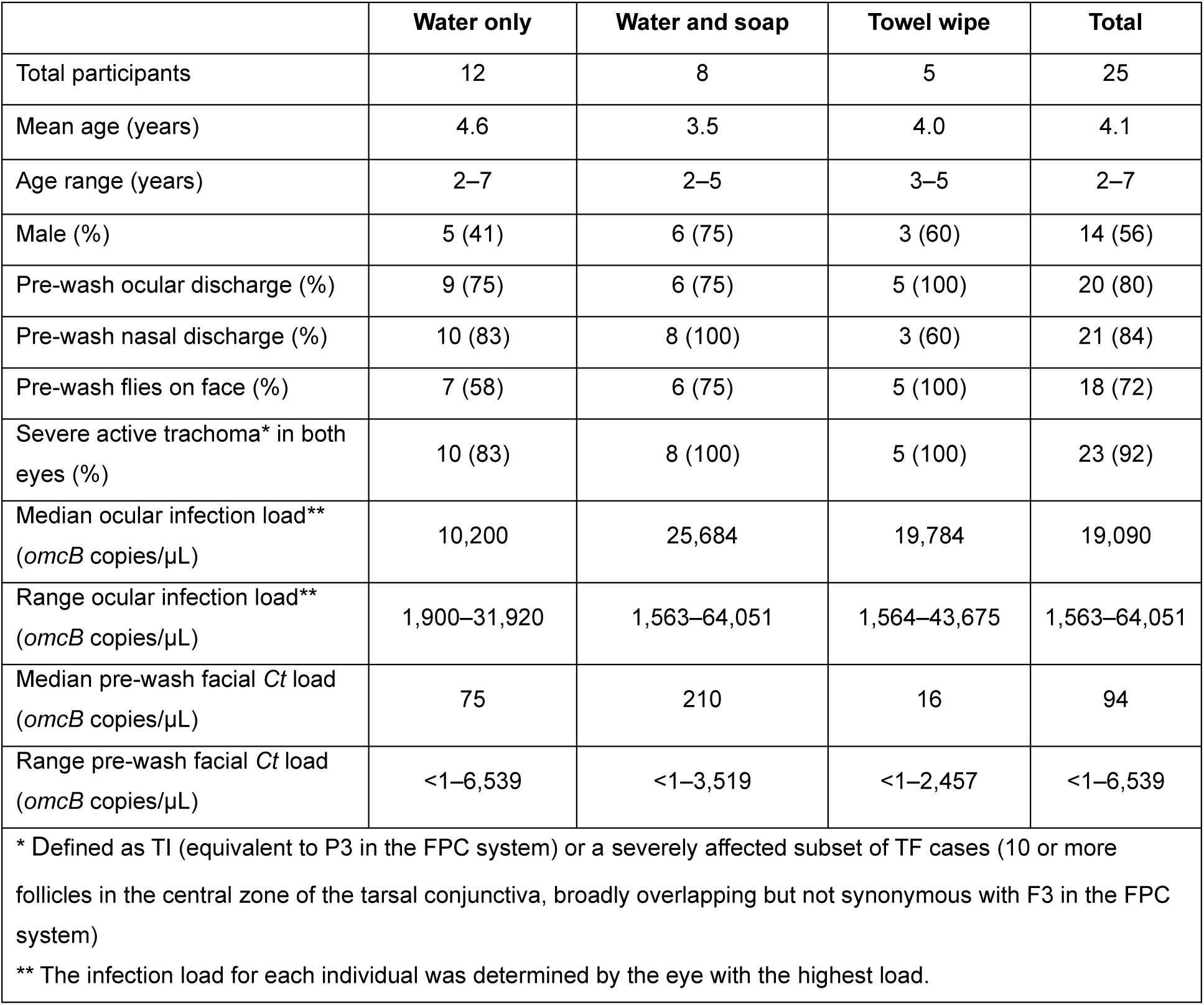
Characteristics of clinical trial participants included in the primary analysis.

### Post-wash removal of Ct and discharge from faces of children by different wash protocols

Washing did not entirely remove *Ct* from any study participant; all 25 (100%) participants across all three arms still had *Ct* on their faces immediately after their assigned washing protocol (Figure 2A). The median pre-wash facial *Ct* load for the primary analysis group reduced from 210 *omcB* copies/µL (range: <1–6,539) to 43 *omcB* copies/µL (range: <1–498) post-wash overall (Mann Whitney U: p=0.006).

**Figure 2.**
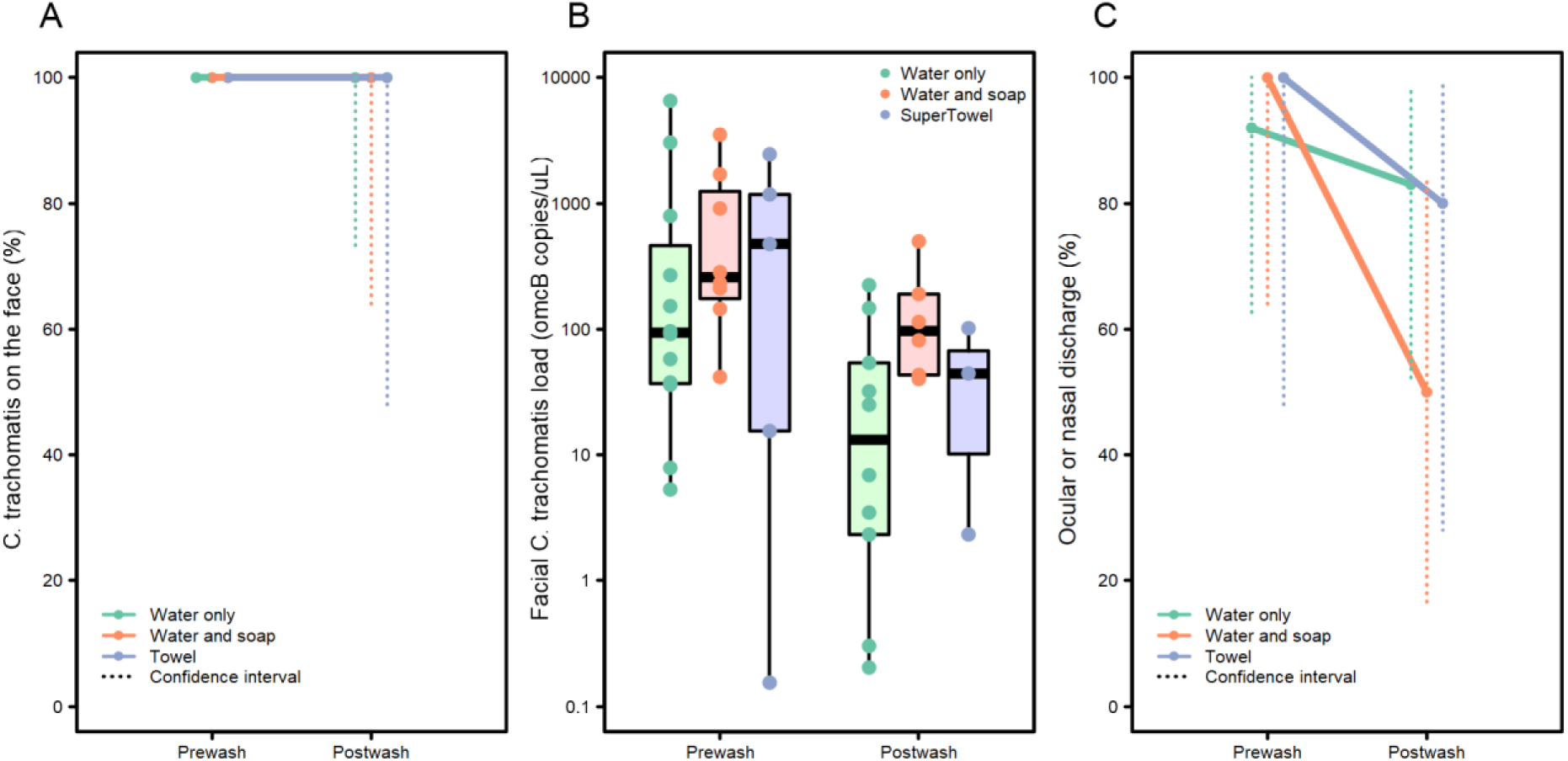
Effect of face-washing protocols on **(A)** the proportion of children with *Chlamydia trachomatis (Ct)* on their faces, (**B**) *Ct* load on children’s faces, and (**C**) the proportion of children exhibiting ocular or nasal discharge, as agreed by two graders in the field.

While there was some evidence of a median *Ct* load reduction within the water-only arm (94 to 15 *omcB* copies/µL; p=0.016), this reduction was not observed in the water-and-soap (257 to 98 *omcB* copies/µL; p = 0.081) or towel-wipe (474 to 44 *omcB* copies/µL; p=0.571) arms.

The median reduction in facial *Ct* load after washing was 65, 289, and 1,132 *omcB* copies/µL for the water-only, water-and-soap and towel-wipe arms, respectively. However, pairwise comparisons revealed no significant difference in effectiveness between any of the protocols in reducing facial *Ct* load in those with conjunctival infection (water-only vs. water-and-soap: p=0.718; water-only vs. towel-wipe: p=0.161; water-and-soap vs. towel-wipe: p=0.548) (Figure 2B).

Ten individuals had *Ct* on their face but no conjunctival infection, thus they were excluded from the primary analysis. Notably, their facial *Ct* load (n=10, median: <1 *omcB* copy/µL, range: <1–531) was significantly lower than in individuals with conjunctival infection (n=25, median: 210 *omcB* copies/µL; range: <1–6539; Mann Whitney U: p<0.001). For these 10 individuals, washing successfully removed all facial *Ct* when assessed immediately post-wash (water-only: 5; water-and-soap: 3; towel-wipe: 2). None of these 10 individuals lived in households with another child with conjunctival infection, though seven lived in *kebeles* with at least one enrolled child with conjunctival infection.

Of participants in the primary analysis, 24 of 25 had visible ocular and/or nasal discharge pre-wash, which was removed in six cases. The one individual without pre-wash discharge also had none post-wash.

Facial cleanliness, assessed in the field, improved with washing: pre-wash, 11/12 (water-only), 8/8 (water-and-soap), and 5/5 (towel-wipe) children had visible discharge. Complete removal of discharge occurred in 1/11 (9%; 95% confidence interval [CI]: <1–41%) with water-only; 4/8 (50%; 95% CI: 16–84%) with water-and-soap; and 1/5 (20%; 95% CI: 1–72%) with a towel. There was no evidence that any protocol was superior (Fisher’s exact test: water-only vs. water-and-soap p=0.109; water-only vs. towel-wipe p=0.515 and water-and-soap vs. towel-wipe p=0.565) (Figure 2A).

Face photograph review suggested improved facial cleanliness, even when discharge remained (Figure 3). Side-by-side comparison showed 21 of 25 (84%) faces were cleaner post-wash, including 16 with visible post-wash discharge. One (4%) face was cleaner pre-wash, and three (12%) were judged unchanged. Graders found 11/12 (92%), 6/8 (75%) and 4/5 (80%) of faces cleaner post-wash in the water-only, water-and-soap and towel-wipe arms, respectively.

**Figure 3.** Selected examples of pre- and post-wash facial photographs showing changes in oculo-nasal discharge and facial cleanliness in children who presented with discharge both before and after washing. (**A**) female, washed with the water-only protocol. (**B**) male, washed with the water-and-soap protocol. (**C**) male, washed with the towel-wipe protocol.

### Impact of face-washing on Ct carriage on the hands of children and their caregivers

*Ct* was detected on the hands of 27 children, including all 25 in the primary analysis group (with conjunctival infection and facial *Ct*) and two additional children with conjunctival *Ct* infection but no facial *Ct.* Twenty-two caregivers also had *Ct* on their hands, all associated with participants with both conjunctival and facial *Ct*.

Of the 27 children with *Ct* on their hands pre-wash, 24 still had it post-wash. Among these, there was no strong evidence for a reduction in load (Mann Whitney U: p=0.074). *Ct* was removed from 1/12 in the water-only arm, 1/10 in the water-and-soap arm and 1/5 in the towel-wipe arm. No evidence of a difference between arms was found (all Fisher’s exact tests: p>0.9)

Of the 22 caregivers with *Ct* on their hands pre-wash, 19 retained it post-wash. One caregiver without *Ct* on their hands pre-wash acquired *Ct* after washing their child’s face with water only. Among those where *Ct* was not removed, there was no evidence for a load reduction (Mann Whitney U: p=0.682). *Ct* was removed from 0/8 of caregivers’ hands in the water-only arm, 2/9 in the water-and-soap arm and 1/5 in the towel-wipe arm. No strong evidence of a difference between arms was observed (all Fisher’s exact tests: p>0.3).

### Persistence of Ct on hands and faces of children and their caregivers for eight hours after washing

Participants in the primary analysis group (with baseline facial and hand *Ct*) retained *Ct* on their hands and faces at all follow-up visits up to 8 hours. Facial *Ct* was transiently absent in one child at 1 hour, and four at 2 hours post-wash. By 4 hours, all had regained facial *Ct* for the study duration (Table 2). Caregivers in this group reported no child face-washing during this period.

**Table 2.** *Chlamydia trachomatis (Ct)* on children’s faces by conjunctival *Ct* infection and pre-wash facial *Ct* status.

| Conjunctival *Ct* infection | Pre-wash facial *Ct* | Number of children with *Ct* on the face (n/N, %) |  |  |  |  |  |  |
| --- | --- | --- | --- | --- | --- | --- | --- | --- |
|  |  | Pre-wash | 0 hour post-wash | 1 hour post-wash | 2 hours post-wash | 4 hours post-wash | 6 hours post-wash | 8 hours post-wash |
| Positive * | Positive * | 25/25 (100) | 25/25 (100) | 24/25 (96) | 21/25 (84) | 25/25 (100) | 25/25 (100) | 25/25 (100) |
| Positive | Negative | 0/8 (0) | 1/8 (13) | 1/8 (13) | 1/8 (13) | 1/7 (14) | 1/8 (13) | 1/8 (13) |
| Negative | Positive | 10/10 (100) | 0/10 (0) | 0/10 (0) | 0/10 (0) | 0/10 (0) | 0/10 (0) | 1/10 (10) |
| Negative | Negative | 0/10 (0) | 0/10 (0) | 0/10 (0) | 0/10 (0) | 0/10 (0) | 0/10 (0) | 0/10 (0) |
| * All three primary trial analyses arms, combined |  |  |  |  |  |  |  |  |

Conversely, all *Ct* was removed from faces in those with pre-wash facial *Ct* but no conjunctival infection. Where facial *Ct* was cleared, it only returned in one individual at eight hours post-wash (Table 2).

Most children with conjunctival infection but no facial *Ct* post-wash remained clear for eight hours. One child with conjunctival *Ct* acquired facial post-wash *Ct*, remaining positive for eight hours (Table 2). No *Ct* was detected on faces of children with neither conjunctival infection nor facial *Ct* at any follow-up timepoint (Table 2).

One caregiver (child: conjunctival negative, facial *Ct* positive) reported washing their child’s face between the 4-hour and 6-hour visits. All other caregivers reported no face-washing between follow-up visits.

The *Ct* load on children’s faces decreased over the first two hours post-wash, then remained relatively stable until 8 hours (Figure 4A). A similar pattern was observed with *Ct* load from infected children’s hands (Figure 4B) and, to a lesser extent, caregivers’ hands (Figure 4C). Hand and face load measurements were highly variable throughout the timeseries.

**Figure 4.**
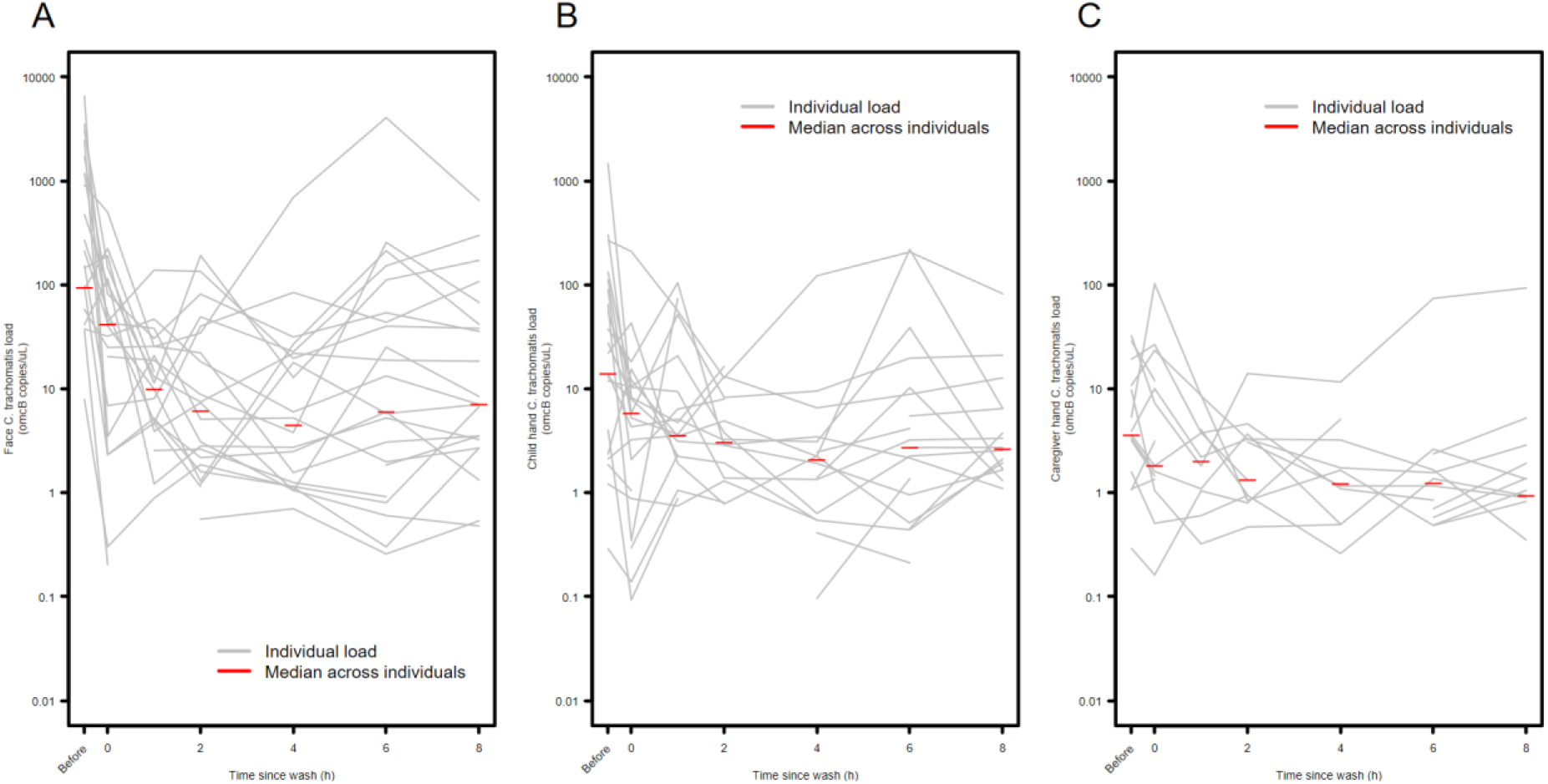
*Chlamydia trachomatis* load recovered from (**A**) children’s faces, (**B**) children’s hands and (**C**) caregivers’ hands over time, after face-washing in a trial comparing three different wash protocols.

## Discussion

Despite successfully removing discharge, a single face wash (water-only, water-and-soap or towel-wipe) did not effectively eliminate *Ct* from the faces of children with conjunctival *Ct* infection and severe active trachoma. The small primary analysis sample size limited our ability to determine protocol superiority. Notably, *Ct* was consistently detected on some children’s faces throughout, while others with severe active trachoma and conjunctival infection had no detectable facial *Ct* at any timepoint, raising questions about whether all high-load/severe inflammation children contribute equally to infection spread.

The finding that face-washing did not remove *Ct* DNA from children’s faces may seem inconsistent with current theories for the SAFE strategy’s F component. However, the washing methods used, even with soap or a damp towel, are unlikely to remove DNA contamination in the way potent laboratory decontamination strategies would (36). Thus, *Ct* DNA persistence post-washing may not be surprising. Furthermore, the clinical significance of residual *Ct* DNA for transmission is unclear. This study did not evaluate the relationship between *Ct* DNA load and pathogen viability (37); therefore, a reduction in DNA load could still imply decreased transmission risk.

Some evidence of washing’s impact on lower *Ct* loads on skin was observed (Table 2, Figure 4), particularly on faces of children without conjunctival *Ct* infection. Our pilot study (Czerniewska *et al.*(16)) also found *Ct* removal from five of 13 children’s faces. While pre-wash facial *Ct* load was not quantified in the pilot, its broader inclusion of TF cases likely yielded a wider range of conjunctival infection loads than our current study, which focused on severe cases. These findings suggest face-washing may be more effective at removing *Ct* in lower*-*load cases.

A reduction in *Ct* load on faces and, to a lesser extent, participants’ hands was observed over the first two hours post-wash, followed by a plateau. Reasons are unclear, but this is not likely to be a prolonged cleaning effect; increased hygiene awareness might have led to more frequent/thorough face cleaning, though most caregivers reported no face washes between visits (although we recognise the potential for self-report bias). Alternatively, ocular discharge and pathogen accumulation overnight might be cleaned off during morning routines, aligning with observations by Harding-Esch *et al.* (19), that morning examinations usually showed cleaner faces. This implies early morning face-washing interventions could be impactful for transmission reduction. The presence of *Ct* on children’s and caregivers’ hands highlights fingers as mechanical vectors. Modest removal from hands after washing suggests dedicated handwashing promotion is needed to target this potential transmission route.

This study had several limitations. Most importantly, it aimed to recruit 141 children with conjunctival and facial *Ct*, but found only 25 (<20% of target), significantly limiting statistical power. Financial and logistical constraints prevented pre-enrolment infection testing; enrolling only severe conjunctival inflammation cases attempted to mitigate the well-documented disconnect between TF and ocular *Ct* infection (37). Based on some previous studies (9,32) and the Stronger SAFE trial in which this study was nested (31), a much higher proportion of conjunctival *Ct* infection was predicted among severe trachoma cases. The analytical limit of detection of the qPCR assay is <10 copies/reaction (35) and this concentration was reproducibly detected in the standard curves run on each plate, therefore this absence of infection is unlikely to be a result of diagnostic failure. However, there is a growing body of literature showing low *Ct* prevalence despite moderate-to-high TF (38–40), e.g., 0.3% ocular *Ct* infection in Amhara Region *woredas* with TF rebounding to >5% (41), 1.3% prevalence of ocular Ct infection in Solomon Islands with TF prevalence of 25.7% (38), 6.7% of F3 cases positive for ocular *Ct* infection in a study in Tanzania (39). Programmatic TF data alone may be insufficient to identify high infection burden sites for future trials. Exclusively studying a minority of severely affected individuals might have selected a population with the highest pathogen loads (42), making *Ct* removal more challenging. Finally, study team presence occasionally induced crying, potentially affecting post-wash assessments of discharge.

Several research priorities emerge. First, defining technical parameters for effective *Ct* removal from infected children’s faces is needed, possibly involving more thorough/regular face-washing, or products to enhance removal of infectious material. Such studies should be co-designed with trachoma-endemic communities for feasibility and acceptability. Second, investigating intra-eye spread routes continues to be important for understanding effective transmission reduction. Third, operational research on the cost and impact of novel face-washing interventions is needed for cost effectiveness.

### Conclusions

While no firm evidence supported a specific face-washing regime for removing *Ct* from faces of children with severe trachoma, this study provides novel insights into the dynamics of facial *Ct* carriage and facial cleanliness in children with trachoma throughout the day. A significant unmet need remains to improve the evidence base for how face-washing can contribute to trachoma elimination.

## Data Availability

The datasets generated and/or analysed during the current study are available in the LSHTM data repository. Anonymised data will be made available upon request for use in ethically-approved research from.

## Declarations

### Ethics approval and consent to participate

The trial was registered with the ISRCTN registry in April 2023 (ISRCTN12814010). The trial was approved by the London School of Hygiene & Tropical Medicine Interventional Ethics Board (16470), the Ethiopia Food and Drug Authority (02/25/33/36), the Ethiopia National Research Ethics Review Committee (03/246/958/22) and the Oromia Regional Health Bureau (BFO/1BTH/1.16/7017).

A parent or guardian of each trial participant was informed of the nature and requirements of taking part and was required to provide written consent on behalf of their child before enrolment. Caregivers were also required to provide written consent to their hands being swabbed.

### Consent for publication

Explicit written approval was obtained for the publication of face photos in trial outputs.

### Availability of data and materials

The datasets generated and/or analysed during the current study are available in the LSHTM data repository at [PERSISTENT WEB LINK TO DATASET]. Anonymised data will be made available upon request for use in ethically-approved research from.

## Acknowledgements

We thank Olika Fekadu, Mekdes Sisay, Mesfin Negewo, Aschlew Abebe, Tsegaye Dechu, and Mestawet Kefeni from the Adama Public Health Referral and Reference Laboratory who processed the specimens. We thank Eva Chong from the London School of Hygiene & Tropical Medicine for administrative support. We thank Chrissy Roberts from London School of Hygiene & Tropical Medicine for writing the python script to randomize face photos for grading.

